# Molecular characterization of hepatitis B virus genotype D and recombinant strains among inmates and blood donors in Northeastern Kenya

**DOI:** 10.1101/2025.10.31.25339225

**Authors:** Vincent Bahati Odallo, Okoti P. Aluora, Wallace Bulimo, George Gachara

**Author notes:** Corresponding author Email - (GG).

## Abstract

**Introduction:** Hepatitis B virus (HBV) persists as a major global public health burden, with hyperendemic prevalence in sub-Saharan Africa. Populations with elevated exposure to percutaneous transmission risks—including incarcerated individuals and healthcare workers—demonstrate heightened HBV susceptibility. Despite this, genomic data from Northeastern Kenya and Kenyan prison populations remain scarce.

**Objective:** To characterize HBV genotypic diversity circulating in Northeastern Kenya, among low risk (blood donors) and high-risk (prison inmates) populations.

**Methods:** A cross-sectional investigation compared HBV seroprevalence and genotypes between incarcerated individuals (n = 130) and voluntary blood donors (n = 130) in Garissa County, Northeastern Kenya. Sample size was calculated using Casagrande’s formula for binomial comparison powered at 80% (α=0.05) to detect a ≥2-fold seroprevalence difference between cohorts. Serum samples underwent Hepatitis B surface antigen (HBsAg) screening, PCR amplification of a 940-bp overlapping surface/polymerase gene, sequencing, and phylogenetic/recombination analyses of the resulting sequences.

**Results:** Seroprevalence was higher among incarcerated individuals (5.4%, 7/130) than blood donors (3.1%, 4/130). HBV DNA was detected in 22 samples. Genotype D dominated both cohorts (81.8%), while genotype A subgenotype A1 occurred exclusively in incarcerated participants (18.2%). All genotype D strains were recombinants: D/A (61%) and D/E (39%). Sequences are accessible in GenBank (accession numbers: PV816552–PV816573).

**Conclusions:** This first genomic study of HBV in Kenyan prisons confirms incarcerated populations as high-risk. The predominance of genotype D—a novel finding in this region—and high recombinant frequency (100% of genotype D strains) underscore significant viral evolution. Expanded genomic surveillance is imperative to define HBV diversity, inform vaccine efficacy monitoring, and optimize control strategies in Northeastern Kenya.

## Introduction

Hepatitis B virus (HBV) remains a major global health burden, with an estimated 254 million individuals living with chronic infection in 2022 and an estimated 1.1 million annual deaths primarily due to liver cirrhosis and hepatocellular carcinoma (1). The virus is genetically diverse and is classified into ten genotypes (A–J) with distinct geographic distributions and clinical implications (2–5). Sub-Saharan Africa, including Kenya, has been identified as a region with a high HBV burden, predominantly characterized by genotypes A, D, and E (6–9). These genotypes have shown a propensity for intergenotypic recombination, a process that plays a critical role in the virus’s evolutionary dynamics, enhancing its adaptability, transmission, and persistence within populations (10–12).

HBV recombination is not merely a molecular curiosity but has significant implications for public health. Recombination events can alter viral fitness, impact the course of disease, and create challenges in diagnosis, treatment, and prevention. Previous studies in Africa have reported the circulation of HBV recombinants involving genotypes A, D, and E, suggesting that these strains are not only evolving but also influencing regional epidemiology (11). In Kenya, earlier research has identified recombinant strains, such as D/E recombinants, but comprehensive molecular data on recombination patterns and their clinical consequences remain limited (8).

Makhoha et al., 2023 recently published a systematic review and meta-analysis of the estimates of HBV infection in Kenya and obtained an overall pooled prevalence estimate of 7.8% (13). Available studies have traditionally surveyed the general Kenyan population, as well as specific populations such as blood donors, HIV patients, pregnant women, and healthcare workers. Geographically the studies have ignored the northern and northeastern regions of the country (14) where anecdotal reports suggest a higher HBV burden. Additionally, current studies have ignored the incarcerated population which is a known HBV high risk group. Consequently, the exact picture of the HBV burden in the country is not fully understood and may be underestimated.

HBV genotypes influence disease progression, treatment response, and likelihood of mutations conferring vaccine escape or antiviral resistance (15). Genotype D has been associated with more severe liver disease in some populations. However, its epidemiological and clinical implications in Kenya remain poorly defined. While previous studies have identified genotypes A, D, and E in Kenya (7, 16), the distribution and impact of recombinant HBV strains are under-reported. This study sought to address these gaps by conducting a molecular analysis of HBV in a group of inmates and voluntary blood donors in the northeastern part of Kenya.

The study population therefore constituted two groups with unique epidemiological characteristics relevant to HBV transmission and molecular diversity. Prison inmates are often at higher risk of HBV infection due to shared living spaces, limited access to healthcare, and behaviours such as drug use or unsafe tattooing practices. Conversely, voluntary blood donors represent a low-risk population routinely screened for infectious diseases, offering valuable insights into HBV strains circulating within the general population. The primary aim of the study was to characterize the burden and genotypic diversity of HBV strains circulating within this neglected population. The findings are expected to provide a clearer picture of HBV burden in the country which is important in developing targeted interventions to curb HBV transmission and progression in line with the global agenda to eliminate viral hepatitis by 2030.

## Materials and methods

### Study Setting

The study was conducted in Garissa County in two study sites namely Garissa County Referral Hospital (GCRH) and Garissa Main Prison both located in Garissa County within the Township area. This County is found to the Northeastern part of Kenya. GCRH receives patients from Garissa County and also neighbouring counties such as Kitui, Tana River and Wajir. Equally, Garissa Main Prison is a host to in-mates from both Garissa County and other parts of the country.

### Study Population

This involved two groups; Voluntary healthy blood donors and Prison inmates. The blood donors presented at Garissa County Referral Hospital (GCRH) while the inmates were incarcerated at Garissa Main Prison. Irrespective of gender and other socio-political characteristics, all voluntary blood donors were selected according to the Kenya National Blood Transfusion Services (KNBTS) criteria. Each consenting donor was aged between 18 – 65 years and weighed at least 50 Kgs. Every consenting inmate aged 18 years and above was also included.

Sample size was determined based on an estimated HBV prevalence of 5% in high-risk populations (prison inmates) and 3% among low-risk populations (blood donors), using standard sample size formula for comparing two proportions (Casagrande et al., 1978) powered at 80% (α=0.05) to detect a ≥2-fold seroprevalence difference between cohorts.

### Ethical considerations

Ethical approval was sought and granted from Kenyatta University Ethics and Review Committee (KU-ERC, application number PKU/2043/I1190). A research permit from the National Commission for Science, Technology & Innovation (NACOSTI) to conduct the research was obtained under License No. NACOSTI/P/20/4150. To further facilitate the study, clearance was sought and obtained from the Kenya National Blood Transfusion Services (KNBTS) trainings board and the Commissioner General and the Trainings Board of the Kenya Prisons Service. Each one of the study participants gave informed consent after the study protocol was explained to them in writing and participated without coercion or remuneration.

### Sample collection and processing

The consenting participants presented themselves for blood collection. Sample collection for the inmates was done at Garissa Main Prison dispensary located within the prison compound while for the blood donors, the collection was done at GCRH Blood Transfusion Unit (BTU) between 03/08/2020 and 31/12/2020. Approximately 4ml of blood was collected from a selected vein of each individual into a plain vacutainer. For the blood donors, after the blood bag was disconnected from the donor, 4 mls of blood from the blood bag was tapped off from each participating donor into a vacutainer. Serum for serological work was prepared by centrifuging whole blood at 3000xg for five minutes. Serum preparation and serological testing was done on site at Garissa Main prison dispensary laboratory for the inmates and at GCRH laboratory for the blood donors. Serum was aseptically aspirated and dispensed into already prepared cryotubes, packaged in a cooler box and transported to Kenyatta University (KU) laboratories where they were refrigerated at −80°C awaiting further processing.

### Serological testing

All the serum samples were screened for the hepatitis B virus surface antigen (HBsAg) using a rapid immunochromatographic test cassette (Amitech Diagnostics Inc.) following manufacturer’s instructions.

### DNA extraction

The HBV DNA was extracted using the Isolate II Genomic DNA Extraction kit (Bioline, Germany) according to manufacturer’s instructions. The eluted DNA was then stored at −20°C awaiting amplification process.

### PCR and sequencing

The overlapping portion of the HBV P and S genes was amplified using a heminested PCR protocol. The PCR amplification reaction was performed in a final volume of 25 μl using sets of primers and protocols described previously (17) to amplify the region 251-1190 from the EcoR1 site of the viral genome.

The first round of PCR reaction contained 0.5µl My Taq DNA polymerase, 10 µl 5X My Taq Reaction buffer, 0.5µl each of 20µM first round primers (251f and 1797r), 7.5µl of nuclease free water and 6µl of the DNA template. Both initial denaturation and denaturation were performed at 95°C for all the samples. An initial denaturation for 2 minutes was followed by 35 cycles of denaturation for 15s, annealing at 58°C for 30s and extension at 72 °C for 30s. A final extension was performed at 72 °C for 2 mins while a final hold was set at 4 °C to terminate the reaction. The second round PCR reaction was performed using 5 µl of the first round PCR reaction product as template while utilizing the second set of primers (251f and 1190r) under the same reaction conditions. Each run utilized negative and positive controls.

The PCR products were visualized and resolved in a 1.5% agarose gel electrophoresis stained with 5µl ethidium bromide (EtBr) and examined using a UV transilluminator. Purification of the HBV PCR positive amplicons before sequencing was done by treating with ExoSAP-IT™ (ThermoFisher Scientific, CA, USA). The purified PCR products were then sequences by bidirectional Sanger sequencing on an ABI 3500xL (Applied Biosystems, CA, USA) sequencer.

### Data Analysis

The resulting forward and reverse reactions were assembled into contiguous nucleotide sequences and manually edited in BioEdit version 7.25 (18) sequence editor. Clustal W program implemented in BioEdit was used in alignment of the resulting nucleotide sequences. The geno2Pheno database (https://hbv.geno2pheno.org) and phylogenetic trees were used for HBV genotypic determination. MrBayes program version 3.1.2 was used for phylogenetic reconstruction (19). First, genotype specific sequences were downloaded from Genbank and the resulting fasta file which included the study sequences was converted into the nexus format using Concatenator (http://cobig2.fc.ul.pt.). The nexus file was then run for 1 million generations aimed at achieving inference of the Bayesian tree. Visualization of the resulting phylogenetic tree and determination of the HBV genotypes was done using Fig Tree version 1.3.1 (http://tree.bio.ed.ac.uk/software/figtree/).

Recombination was analysed for sequences whose sub-genotype identity could not be resolved by phylogenetic reconstruction using the HBV NCBI genotyping tool (https://www.ncbi.nlm.nih.gov/projects/genotyping/formpage.cgi).

## Results

### Seroprevalence of HBV

A total of 260 study participants were enrolled in the current study, out of whom 130 were voluntary blood donors and 130 were inmates. Among the blood donors, 4/130 (3.1%) were HBsAg seropositive while among the inmates, 7/130 (5.4%) were HBsAg seropositive as shown in Table 1. Although seroprevalence was higher among inmates (5.4%) than blood donors (3.1%), this difference did not reach statistical significance (p=0.31, chi-square test)

**Table 1.** HBV seroprevalence among blood donors and inmates.

| **HBV**<br>**Seropositivity** | **Blood Donors** |  | **Inmates** |  |
| --- | --- | --- | --- | --- |
|  | Frequency | Percent (%) | Frequency | Percent (%) |
| **Negative** | 126 | 96.9 | 123 | 94.6 |
| **Positive** | 4 | 3.1 | 7 | 5.4 |
| **Total** | **130** | **100.0** | **130** | **100.0** |

### Prevalent HBV genotypes

A 940bp HBV DNA was successfully amplified and sequenced from twenty-two (22) samples which included several seronegative samples (Gene bank accession numbers: PV816552-PV816573). The geno2Pheno database showed that out of the 22 HBV sequences, 18 (81.8%) were identified as genotype D sub-genotype D4 among both blood donor and inmate populations. Genotype A sub genotype A1 was identified in 4 (18.2%) sequences which were all derived from inmates as shown in Table 2 below.

**Table 2:**
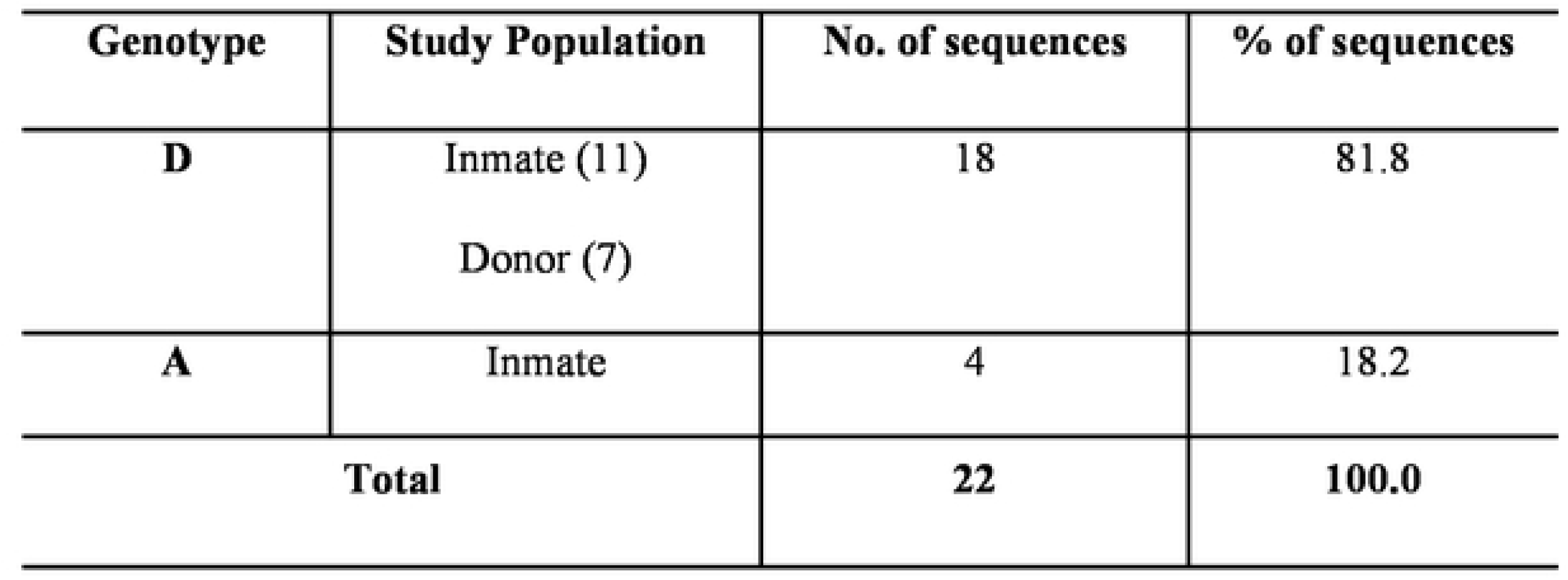
HBV Genotypes among the study participants.

The phylogenetic analysis confirmed the identities of the circulating HBV genotypes and also the A1 subgenotype. However, the sub-genotype identity of the identified genotype D could not be resolved. The genotype D sequences from this study did not cluster with published subgenotype D4 sequences but instead distinctly clustered away from the other sub-genotypes. This cluster of local genotype D sequences from the study was well supported with a posterior probability of 91% as shown in Fig 1.

**Fig 1:**
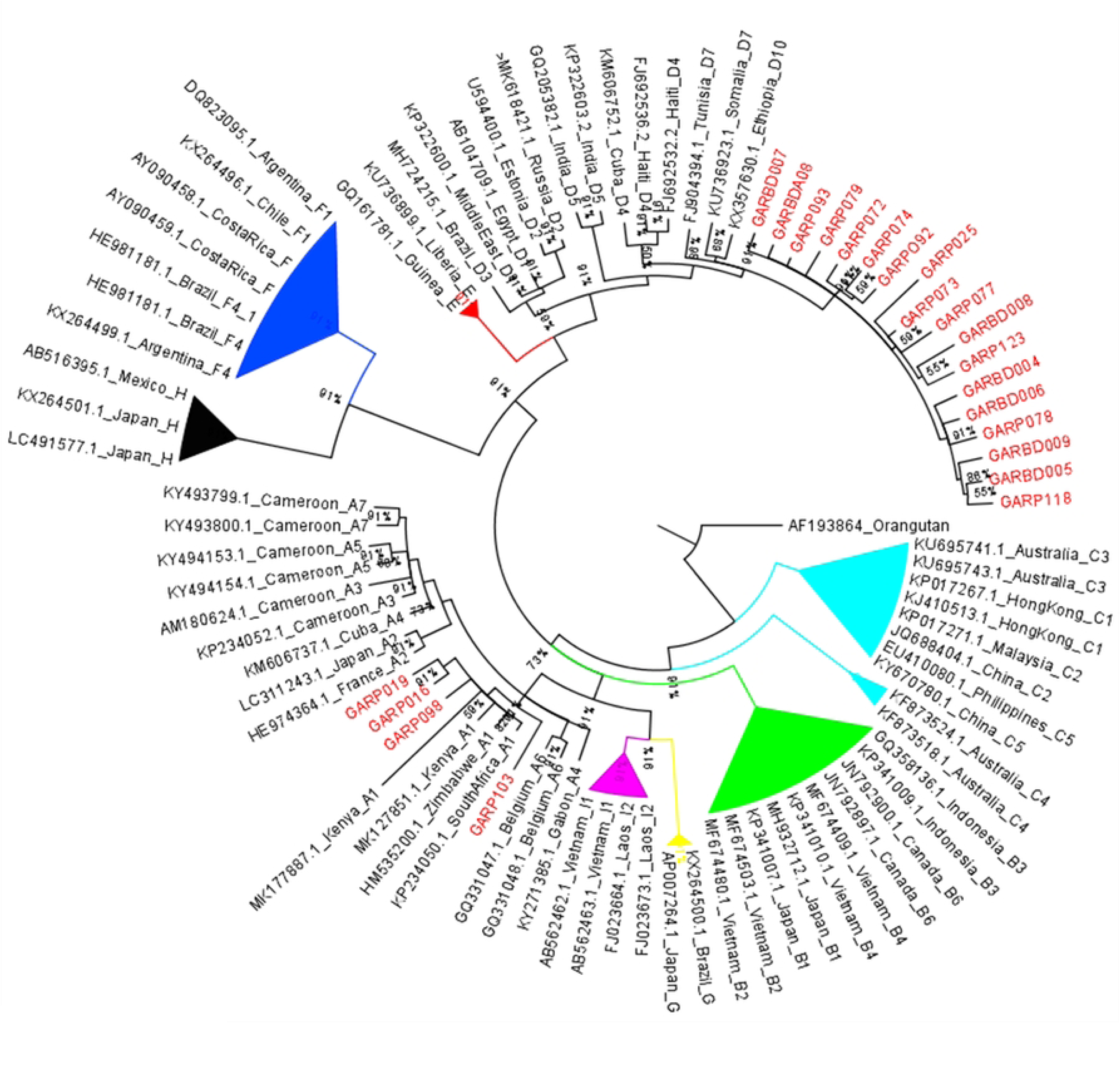
Phylogenetic Tree of HBV sequences from this study alongside published sequences. Sequences from this study are marked in red at the taxa and clustered with Genotype A and D.

### HBV genotype D recombinants

The 18 genotype D sequences were checked for recombination in an attempt to resolve their sub-genotype identity. All the sequences were found to be recombinants with genotype A and E. There were 11/18 genotype D/A recombinants and 7/18 genotype D/E recombinants as shown in supplementary figures S1 and S2.

## Discussion

The magnitude of the heavy toll resulting from Hepatitis on lives, societies and health systems globally is encapsulated in the 2030 Agenda for Sustainable Development (20). Specifically, Sustainable Development Goal 3 (SDG 3) aims to “Ensure healthy lives and promote well-being for all at all ages” with target 3.3 aiming to end the epidemics of AIDS, tuberculosis, malaria, and neglected tropical diseases and combat hepatitis, waterborne and other communicable diseases by 2030. In view of this target, the WHO has set the goal of eliminating viral hepatitis as a major public health threat by 2030. A fundamental strategy to achieve this target calls for countries to understand the populations most affected. In Kenya, there is scanty information on hepatitis infection among prison inmates and the populations in the Northern region (14) thus necessitating this study.

The seroprevalence of HBV in the two study populations was observed to differ. Inmates who are classified as high risk for HBV infection had a HBsAg seropositivity of 5.4% compared to 3.1% among the blood donors who are classified as low risk. These findings are in agreement with a recent systematic review of HBV prevalence in Kenya using 23 studies published between January 2000 and December 2021. The review found a pooled HBV prevalence of 3.1% (95% CI 2.62-4.01%) among the low-risk group which includes blood donors. It also found a pooled prevalence of 5.58% (95% CI 3.46-7.7% and, 6.17% (95% CI 4.4-9.94) among moderate risk and high-risk groups respectively (14). The current study therefore affirms the categorization of blood donors as a low-risk group and inmates as a high-risk group in HBV transmission. It is important to note that the systematic review observed that no study had been conducted in Northern Kenya and also among inmates in the country.

The genotypic analysis performed in this study using the Geno2Pheno database identified genotype D, sub-genotype D4 as the dominant HBV genotype, accounting for 81.8% of the sequenced samples across both blood donor and inmate populations. Among the inmate population, a smaller proportion of subjects exhibited genotype A, sub-genotype A1, underscoring the coexistence of multiple genotypes within this cohort. The exclusive detection of sub-genotype A1 among inmates, suggests a possible distinct transmission dynamics within the incarcerated population. Earlier studies in Kenya have documented the circulation of HBV genotypes A, D, and E (7, 16, 21), with genotype A1 consistently identified as the predominant genotype (6, 16). However, the dominance of genotype D, as observed in this study, represents a novel finding, diverging from the established pattern in previous research. Genotype D dominance, as observed here, may impact disease progression and treatment outcomes given evidence of higher risk of liver disease progression associated with certain genotype D sub-genotypes (22).

The distribution of HBV genotypes and sub-genotypes is largely influenced by ethnicity and migration (23). Consequently, the predominance of genotype D in Northeastern Kenya, as reported here, could be attributed to the socio-demographic dynamics of the region. This area is predominantly inhabited by the Somali ethnic group, which constitutes a significant proportion of the population in the Horn of Africa. Studies have suggested that HBV genotype D is widespread in East Africa, including among Somali populations (24, 25). The potential introduction of genotype D into Northeastern Kenya through migration or trade with neighboring regions may have further contributed to its prevalence in this region.

The genotype D sequences in this study revealed unexpected phylogenetic patterns. Despite utilizing published genotype D sub-genotype reference sequences, the local genotype D sequences failed to align with any recognized sub-genotype (D1-D10). Instead, these sequences formed a separate, well-supported cluster (posterior probability = 91%), distinct from established sub-genotypes. The absence of clustering with recognized HBV genotype D sub-genotypes raises intriguing possibilities. One explanation is that the observed sequences represent a new HBV sub-genotype within genotype D, defined by a genome divergence of at least 4%, as established in HBV classification guidelines (24, 26). We could not test this hypothesis since the study did not sequence full genomes. Alternatively, these sequences could represent genotype D recombinants, as recombination events are not uncommon in HBV and have been documented in other studies (11, 12). This hypothesis is further supported by the high genetic diversity within genotype D, as well as reports of recombinant strains globally, including in Africa (8, 11, 27).

Previous studies in Kenya have detected both genotype D sub-genotypes D1 and D4 (16, 21). However, outliers that do not conform to any recognized sub-genotype have also been reported, such as an unclassified HBV genotype detected in 2013 (16). The presence of recombinant strains of HBV, particularly D/A and D/E recombinants, underlines the complexity of viral evolution in the region. The high frequency of D/A recombinants (61%) observed in this study suggests that genotype D may serve as a major parental strain for recombination with other prevalent genotypes in Kenya, particularly genotype A. The remaining recombinant strains being D/E suggest the potential for cross-genotypic interaction between strains of different African lineages. In fact, as observed previously, genotype D is widely distributed across Africa, with both D1 and D4 sub<u>-</u>genotypes circulating in various regions (8). These sub<u>-</u>genotypes could serve as reservoirs for further recombination (28), leading to new viral strains that may possess a different set of genetic characteristics compared to their parental strains.

The identification of these recombinant strains is not only a reflection of the virus’s adaptability but also an indicator of the dynamic viral population in Kenya. This is consistent with earlier studies which suggested that HBV genotypes A, D, and E show significant geographic overlap in regions of Africa with high migration patterns, such as East Africa (28). This migratory behaviour, driven by trade and population movement, could facilitate the exchange of different viral strains across borders, creating opportunities for recombination events.

The high frequency of HBV D/A and D/E recombinant strains observed in this study (81.8% of genotype D cases) underscores the significant genetic plasticity of HBV in Northeastern Kenya. Such recombination between co-circulating genotypes is a recognized evolutionary phenomenon (29) and complicates molecular surveillance by obscuring true transmission networks. Critically, these recombinant forms may impact diagnostic accuracy if commercial assays target recombinant-prone genomic regions and could theoretically alter vaccine efficacy or antiviral response—though clinical implications of D/A recombinants remain uncharacterized and warrant further investigation.

Our findings must be interpreted in light of two limitations. First, HBsAg screening relied on rapid immunochromatographic tests (sensitivity: 90–95%) rather than higher-sensitivity EIAs (>99%) or PCR (1), potentially underestimating true seroprevalence. Second, sequencing of only a 940-bp P/S fragment precluded comprehensive recombination mapping (e.g., breakpoint identification), full sub-genotype characterization, and assessment of drug-resistance mutations. Thus, whole-genome sequencing of HBV-positive samples from this region is essential to resolve recombinant structures, detect emerging variants, and clarify clinical implications.

Notwithstanding its limitations, this study provides the first genomic characterization of hepatitis B virus (HBV) in Kenya’s Northeastern region, revealing two critical findings: a dominant circulation of genotype D among incarcerated populations (representing 81.8% of HBV-DNA positive samples) and blood donors, coupled with the emergence of recombinant strains (D/A: 61%; D/E: 39%) indicative of novel viral variants. These results significantly advance our understanding of HBV’s evolutionary trajectory in East Africa, a region where genotype A historically predominated. The high recombinant frequency signals active viral adaptation with tangible public health implications: diagnostic reliability may be compromised if commercial assays target recombinant-prone genomic regions, while vaccine efficacy could theoretically be undermined through immune-escape mutations, and antiviral treatment efficacy may require reassessment should recombinants alter drug-resistance profiles. To address these challenges, we recommend implementing regionally tailored genomic surveillance programs, validating existing diagnostics against circulating recombinant strains, and prioritizing whole-genome sequencing to resolve sub-genotypic diversity and recombination breakpoints. Sustained monitoring of these evolving strains is essential to optimize prevention and treatment strategies for high-risk populations in Kenya.

## Data Availability

The Sequences from the study are deposited in GenBank under Accession numbers: PV816552-PV816573

## Acknowledgements

The authors would like to extend their sincere gratitude to the study participants for their invaluable contribution and consent to participate in this research. Without their cooperation, this study would not have been possible. We also acknowledge the support of the healthcare staff and institutions involved in data collection and sample processing. Special thanks are given to the laboratory technicians for their meticulous work in sequencing and analyzing the samples.

## Funding

This work was funded by the Authors’ resources.

## Authors’ contributions

VBO, GG, and WB conceptualized the study. VBO and GG collected and performed the laboratory analysis. VBO, GG, WB and POA analyzed the data. VBO, GG and POA prepared the manuscript. GG supervised he study. All authors read and approved the final manuscript.

## Supporting information

**S1** Figure of a recombination analysis of a HBV genotype D sequence showing D/E recombination.

**S2** Figure of a recombination analysis of a HBV genotype D sequence showing D/A recombination.

## Notes

### Competing Interest Statement

The authors have declared no competing interest.

### Funding Statement

The author(s) received no specific funding for this work.

### Author Declarations

Kenyatta University Ethics and Review Committee (KU-ERC)

